# Presence of Live SARS-CoV-2 Virus in Feces of Coronavirus Disease 2019 (COVID-19) Patients: A Rapid Review

**DOI:** 10.1101/2020.06.27.20105429

**Authors:** Prabjot Sehmi, Isaac Cheruiyot

**Affiliations:** School of Medicine, University of Nairobi, Kenya

**Author notes:** **Address all correspondence and reprint requests to:** Isaac Cheruiyot, BS., School of Medicine, University of Nairobi, Nairobi, Kenya.

**Keywords:** COVID-19, SARS-CoV-2, feco-oral transmission, Feces

## Abstract

Coronavirus disease 2019 (COVID-19) is a rapidly escalating pandemic that has spread to many parts of the world. The disease has already affected over 6 million individuals, with over 400,000 fatalities. Recent studies have confirmed the presence of SARS-CoV-2 nucleic acids in feces of Coronavirus disease 2019 (COVID-19) patients using RT-PCR tests. It is however, still unclear as to whether or not live SARS-CoV-2 virus is actually present in feces of these patients. In this rapid review, we systematically analyzed literature to establish any evidence of live SARS-CoV- 2 virus in fecal samples of COVID-19 patients. We identified 4 studies (one case report, 2 case series and 1 cohort study) where the SARS-CoV-2 was successfully isolated from fecal samples of COVID-19 patients using culture techniques. Therefore, there is some evidence COVID-19 could shed live SARS-CoV-2 virus via the gastro-intestinal tract. Larger studies are needed to corroborate these findings, as well as to determine its potential for disease transmission and infection, and possible implications for COVID-19 discharge and isolation policies.

## INTRODUCTION

The coronavirus disease 2019 (COVID-19), which was first reported in Wuhan, China in December 2019, has achieved a pandemic status. As of 8^th^ June 2020, the disease had spread to over 180 countries and territories, with over 6,931,000 cases and >400,857 fatalities.^1^

The disease is thought to be mainly spread through respiratory droplets and by fomites.^2^ Whereas the disease primarily affects the respiratory system, a subset of COVID-19 patients presents with gastro-intestinal symptoms,^3,4^ raising concerns of potential alternative routes of viral shedding and disease transmission. The evidence supporting the potential of feco-oral transmission of COVID-19 has been predominantly based on the detection of viral ribonucleic acid (RNA) in feces using reverse transcriptase polymerase chain reaction (RT-PCR).^5-7^ However, this technique does not usually distinguish infectious from non-infectious nucleic acid.^8^ Therefore, detection of viral RNA in stool may not necessarily imply potential for transmission or infectivity. Stronger evidence of possibility of feco-oral transmission of COVID-19 may however be obtained by isolation and demonstration of live virus in stool.

In this rapid review, we systematically analyze literature to establish if there is any evidence of the presence of live Severe Acute Respiratory Syndrome Coronavirus 2 (SARS-CoV-2) in feces of COVID-19 patients. This may have potential implications for management of COVID-19 patients, as well as discharge and isolation policies.

## METHODS

### Literature Search Strategy

A comprehensive and systematic search of literature from November 1, 2019 to June 6^th^, 2020 was conducted on the Medline (PubMed interface) and China National Knowledge Infrastructure (CNKI) to identify studies eligible for inclusion. The electronic search was carried out using the strategy as follows: ((((SARS-CoV-2) OR (Live Virus)) AND (((Culture) OR (Isolation)) OR (Identification))) AND (((Feces) OR (Stool)) OR (Anal swabs)). No language restriction was applied. When the articles were published by the same study group and there was an overlap of the search period, only the most recent article was included to avoid duplication of data. The PubMed function “related articles” was used to extend the search. Also, we searched major infectious disease and general medicine journals reporting articles about COVID-19 infection to look for additional studies. We then performed hand-search of the bibliography of included studies, to detect other potentially eligible investigations.

### Eligibility Criteria

The search results were screened by title and abstract, with those of potential relevance evaluated by full text. Studies were deemed eligible for inclusion if they fulfilled the following criteria: (1) were case reports/case series/cohort studies, (2) included patients with a reverse transcriptase polymerase chain reaction (RT-PCR)-confirmed COVID-19 diagnosis, (3) demonstrated live SARS-CoV-2 virus in feces of COVID-19 patients.

### Data Extraction

For each study, the following information was extracted: the surname of the first author and the year of publication, the geographical region where the study was performed, study setting, sample size, number of patients tested for live virus in feces, number of patients with live virus in feces, methods of isolation & identification of live virus in feces.

### Synthesis of findings

Due to the limited data available, pooling of data was not performed. Instead, the results were summarized in a “summary of findings table” and a narration provided.

## RESULTS

### Study Identification and Characteristics of the Included Studies

The initial search produced 278 potentially relevant articles. Following primary screening and removal of duplicates, 63 articles were assessed by full text for eligibility. Of these, 59 articles were excluded since they were review articles, commentaries, or other editorial materials, or did not contain data on isolation of live in stool specimens in COVID-19 patients. Therefore, 4 articles were included in the final synthesis (figure 1 and table 1). All studies were from China. Essential characteristics of the included studies are provided in table 1.

**Table 1:** Characteristics of the included studies

| **Study** | **Setting** | **Study Design** | **Sample Size** | **No. of Patients Tested for Live Virus in Feces** | **No of Patients with Live Virus in Feces** | **Methods of Isolation & Identification of Live Virus in Feces** |
| --- | --- | --- | --- | --- | --- | --- |
| **Zhang 2020** | China (Biosafety Level 3 (BSL-3) Laboratory of the National Institute for Viral Disease Control and Prevention) | Case report | 1 | 1 | 1 | Virus isolated using Vero cells, then demonstrated using electron microscopy |
| **Xiao F et al., 2020a** | China (Fifth Affiliated Hospital of Sun Yat-Sen University) | Case series | 28 | 3 | 2 | Virus isolated using Vero cells, then demonstrated using electron microscopy |
| **Xiao F et al., \* 2020b** | China (Fifth Affiliated Hospital, Sun Yat-sen University) | Case series | Not reported | Not reported | Authors mention some positive isolation of live virus in some patients | Not reported |
| **Wang et al., 2020** | China (Hubei, Shandong and Beijing) | Cohort | 205 | 4 | 2 | Culture and electron microscopy |

*\* These authors reported unpublished findings on isolation of live virus*

**Figure 1:**
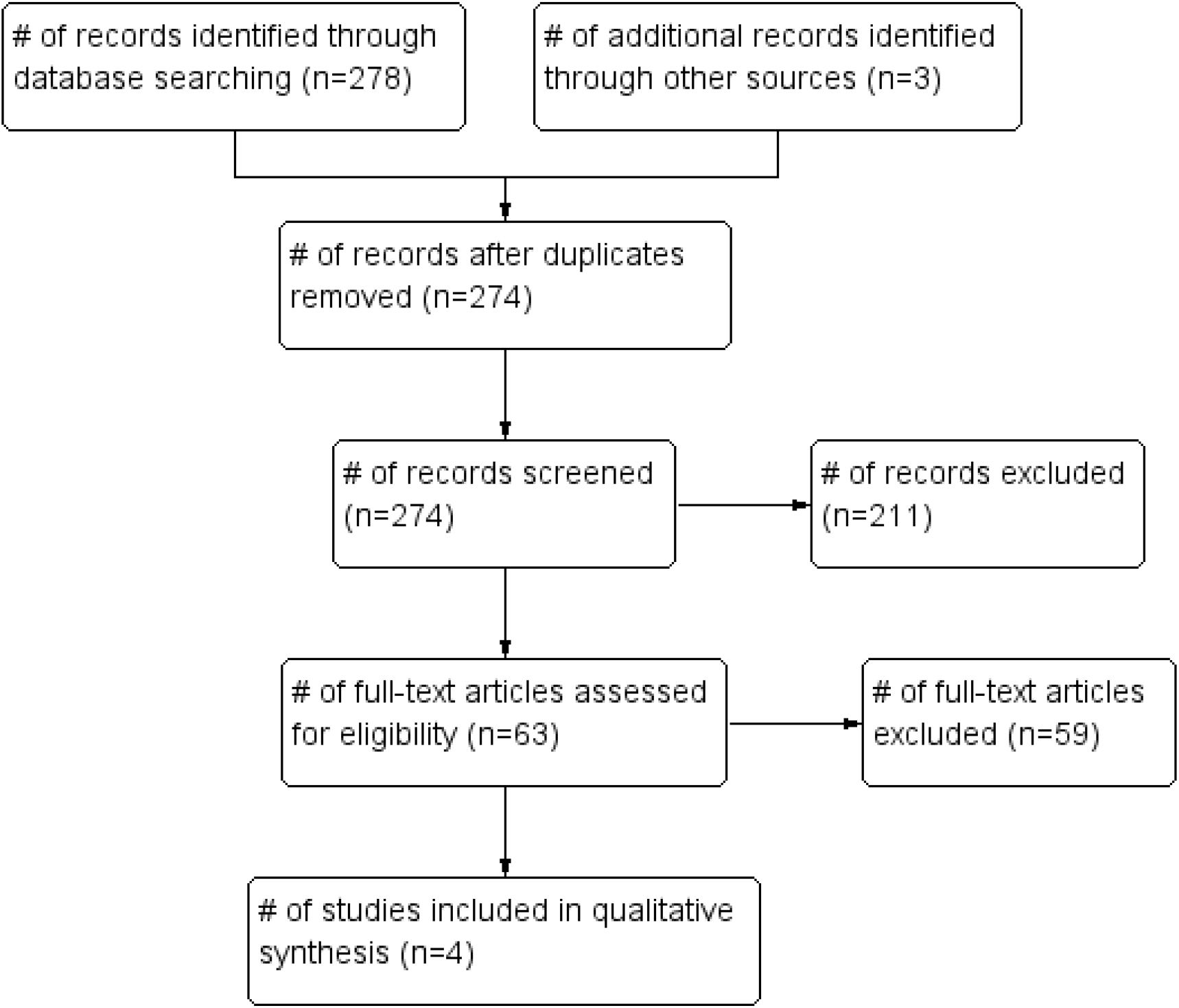
PRISMA flow diagram for the included studies.

### Identification of Live SARS-CoV-2 virus in feces of COVID-19 patients

A total of 4 studies describing isolation of live SARS-CoV-2 virus in fecal samples of COVID- 19 patients were included.^9-12^ The first study was by Zhang and colleagues ^9^ who successfully isolated live SARS-CoV-2 virus in the stool sample of a patient with severe COVID-19 using Vero cell culture and demonstrated it using electron microscopy. Using similar methods, Xiao et al.,^10^ were able to isolate and demonstrate the live virus in 2 out of 3 patients (67%) who had tested positive for SARS-CoV-2 RNA in stool samples tested by RT-PCR. The same authors in another report mentioned their unpublished findings on successful isolation of infectious SARS-CoV-2 from stool.^11^ The fourth study was by Wang and colleagues^12^, conducted across 3 regions in China (Hubei, Shandong and Beijing) on 205 COVID-19 patients. Identification of live virus in stool was attempted successfully in 2 out of 4 patients. It is noteworthy that neither of these patients had gastrointestinal symptoms.

## DISCUSSION

The findings from these four studies, though limited by the small sample sizes, confirm that live SARS-CoV-2 virus is present in fecal samples of COVID-19 patients, and therefore supports the hypothesis that COVID-19 could potentially be transmitted via the feco-oral route.

These findings have important implications. First, isolation of live virus in feces implies that there is a potential risk of transmission of COVID-19 though aerosol generating procedures such as colonoscopy, endoscopy, laparoscopy, electrocautery. Appropriate cautionary measures such as use devices to filter released carbon dioxide during laparoscopy, as well as the use personal protective equipment when performing these procedures. Second, these findings affirm the need for routine testing of fecal samples of COVID-19 patients so as to enable the institution of appropriate public health policies to minimize potential feco-oral transmission of COVID-19. Our study was limited by the small number of the included studies, which were mainly case reports and case series. Larger high-quality studies are urgently needed to better characterize the magnitude of live SARS-CoV-2 viral shedding in feces, as well as its potential for disease transmission and infection, and possible implications for COVID-19 discharge and isolation policies.

In conclusion, there’s some evidence that COVID-19 patients could shed live SARS-CoV-2 virus via feces.

## Data Availability

All supporting data are within the manuscript

## REFERENCES

1. World Health Organization (WHO). Coronavirus disease 2019 (COVID-19) Situation Report – 140. 2020. Accessed 9th June 2020. https://www.who.int/docs/default-source/coronaviruse/situation-reports/20200608-covid-19-sitrep-140.pdf?sfvrsn=2f310900_2

2. Rothan HA, Byrareddy SN. The epidemiology and pathogenesis of coronavirus disease (COVID-19) outbreak. Journal of autoimmunity. 2020 Feb 26:102433.

3. Gu J, Han B, Wang J. COVID-19: gastrointestinal manifestations and potential fecal–oral transmission. Gastroenterology. 2020 May 1;158(6):1518–9.

4. Kotfis K, Skonieczna-Żydecka K. COVID-19: gastrointestinal symptoms and potential sources of 2019-nCoV transmission. Anaesthesiology intensive therapy. 2020 Mar 23;52(1).

5. Chen Y, Chen L, Deng Q, Zhang G, Wu K, Ni L, Yang Y, Liu B, Wang W, Wei C, Yang J. The presence of SARSDCoVD2 RNA in the feces of COVIDD19 patients. Journal of Medical Virology. 2020 Apr 3.

6. Zhang T, Cui X, Zhao X, Wang J, Zheng J, Zheng G, Guo W, Cai C, He S, Xu Y. Detectable SARSDCoVD2 viral RNA in feces of three children during recovery period of COVIDD19 pneumonia. Journal of Medical Virology. 2020 Mar 29.

7. Kipkorir V, Cheruiyot I, Ngure B, Misiani M, Munguti J. Prolonged SARSDCovD2 RNA Detection in Anal/Rectal Swabs and Stool Specimens in COVIDD19 Patients After Negative Conversion in Nasopharyngeal RTDPCR Test. Journal of Medical Virology. 2020 May 13.

8. Atkinson B, Petersen E. SARS-CoV-2 shedding and infectivity. The Lancet. 2020 Apr 25;395(10233):1339–40.

9. Zhang Y, Chen C, Zhu S, Shu C, Wang D, Song J, Song Y, Zhen W, Feng Z, Wu G, Xu J. Isolation of 2019-nCoV from a stool specimen of a laboratory-confirmed case of the coronavirus disease 2019 (COVID-19). China CDC Weekly. 2020 Feb 1;2(8):123–4.

10. Xiao F, Sun J, Xu Y, Li F, Huang X, Li H, et al. Infectious SARS-CoV-2 in feces of patient with severe COVID-19. Emerg Infect Dis. 2020 Aug [date cited]. https://doi.org/10.3201/eid2608.200681

11. Xiao F, Tang M, Zheng X, Liu Y, Li X, Shan H. Evidence for gastrointestinal infection of SARS-CoV-2. Gastroenterology. 2020 May 1;158(6):1831–3.

12. Wang W, Xu Y, Gao R, Lu R, Han K, Wu G, Tan W. Detection of SARS-CoV-2 in different types of clinical specimens. Jama. 2020 May 12;323(18):1843–4.

